# Dysregulated lipid metabolism precedes onset of psychosis in people at clinical high risk

**DOI:** 10.1101/2020.01.30.20019851

**Authors:** Alex M. Dickens, Partho Sen, Matthew J Kempton, Neus Barrantes-Vidal, Conrad Iyegbe, Merete Nordentoft, Thomas Pollak, Anita Riecher-Rössler, Stephan Ruhrmann, Gabriele Sachs, Rodrigo Bressan, Marie-Odile Krebs, Lieuwe de Haan, Mark van der Gaag, Lucia Valmaggia, Tuulia Hyötyläinen, Matej Orešič, Philip McGuire, the EU-GEI High Risk Study

## Abstract

A key clinical challenge in the management of individuals at clinical high risk for psychosis (CHR) is that it is difficult to predict their future clinical outcomes. Here, we investigated if the levels of circulating molecular lipids are related to adverse clinical outcomes in this group. Serum lipidomic analysis was performed in 263 CHR individuals and 51 healthy controls (HC), who were then clinically monitored for up to five years. Machine learning was used to identify lipid profiles that discriminated between CHR subjects and HC, and between subgroups of CHR subjects with distinct clinical outcomes. At baseline, compared to HC, CHR subjects (independent of outcome) had higher levels of triacylglycerols (TGs) with a low acyl carbon number and a double bond count, as well as higher levels of lipids in general. CHR subjects who subsequently developed psychosis (n=50) were distinguished from those that did not (n=213) on the basis of lipid profile at baseline, using a model with an AUC = 0.81 (95% CI = 0.69-0.93). CHR subjects who became psychotic had lower levels of ether phospholipids than CHR individuals who did not (p<0.01). Collectively, these data suggest that lipidomic abnormalities predate the onset of psychosis, and that blood lipidomic measures may be useful in predicting which CHR individuals are most likely to develop psychosis.

## Introduction

A key clinical challenge in the management of individuals at clinical high risk for psychosis (CHR) is that it is difficult to predict their clinical outcomes^1^. Identifying biomarkers that could be used to stratify CHR subjects according to these different outcomes would facilitate more personalized clinical intervention in this group.

Recent studies have shown that metabolic co-morbidities such as weight gain, insulin resistance, altered glucose metabolism and dyslipidemias are common in patients with psychotic disorders^2-4^. Although these can arise as a result of unhealthy lifestyles and treatment with antipsychotic medication^5^, there is growing evidence that they are already present at the onset of psychosis, in patients who are not obese and are medication-naïve^6, 7^.

Changes in the concentrations of specific groups of metabolites, including lipids, are sensitive and specific to several factors that can affect the risk of psychosis, such as genetic variation, environmental exposure, neurodevelopment, age, immune system function, and stress. Metabolomics has therefore emerged as a powerful tool for the characterization of host-environment interactions and complex phenotypes like psychosis^8^. In patients who have recently developed psychosis, rapid weight gain is associated with alterations in circulating lipids that are linked with non-alcoholic fatty liver disease (NAFLD) and insulin resistance^9, 10, 11^. However, the extent to which metabolomics abnormalities are evident in people at Clinical High Risk (CHR) for psychosis, before the onset of illness, has not been investigated before.

We addressed this issue by measuring the levels of circulating molecular lipids in a large sample of CHR individuals, and examining their relationship to the onset of psychosis and other clinical outcomes in this group. We performed comprehensive mass spectrometry (MS)-based lipidomics in serum samples that were collected at baseline from a cohort of CHR individuals. This cohort was then followed up for at least two years to determine their clinical outcomes. We first tested the hypothesis that the CHR group (independent of clinical outcome) would show alterations in metabolomic measures relative to healthy controls. Our second hypothesis was that within the CHR sample, the levels of molecular lipid measures at baseline would be associated with three clinical outcomes at follow up, specifically, persistence of symptoms, transition to psychosis, and a low level of functioning.

## Methods

### Study Population

The EU-GEI (European network of national schizophrenia networks studying Gene-Environment Interactions) High Risk study is a multi-centre longitudinal observation study of individuals at Clinical High Risk (CHR) for Psychosis. A total of 344 CHR individuals identified using Comprehensive Assessment of At-Risk Mental States (CAARMS) criteria^12^ were recruited at eleven sites (Amsterdam, Barcelona, Basel, Cologne, Copenhagen, London, Melbourne, Paris, São Paulo, The Hague, Vienna). Sixty-seven heathy controls were recruited at four of these sites (Amsterdam, London, Melbourne, The Hague). Each site obtained ethical permission for the study and written informed consent was obtained from all participants. CHR and control participants were interviewed at baseline, had repeat assessments at 12 and 24 months, then further clinical follow up for up to five years. Within the CHR group, 65 (18,9%) transitioned to psychosis during the study: 57 within two years and 8 after two years.

#### Inclusion and Exclusion Criteria

Presence of the inclusion criteria for the CHR state were determined using the CAARMS^12^. Exclusion criteria for CHR and controls were past/present diagnosis of a psychotic disorder, inadequate understanding of the language local to the site, or not able or willing to provide a blood or saliva sample. Previous exposure to antipsychotic medication was not an exclusion criterion for CHR subjects, but was for healthy controls.

#### Demographic and Clinical Measures

Data on age, gender, ethnicity, height and weight were obtained using the modified Medical Research Council Sociodemographic Schedule^13^. At baseline and follow-up, trained raters assessed participants using the CAARMS and the Global Assessment of Function (GAF) split scale^14^. Inter-rater reliability (IRR) was assessed using online CAARMS and GAF training videos. CHR participants were determined to be in remission if they no longer met the criteria for the CHR state (assessed using the CAARMS) at follow-up.

Demographic characteristics of the study population are shown in **Table 1**.

**Table 1.**
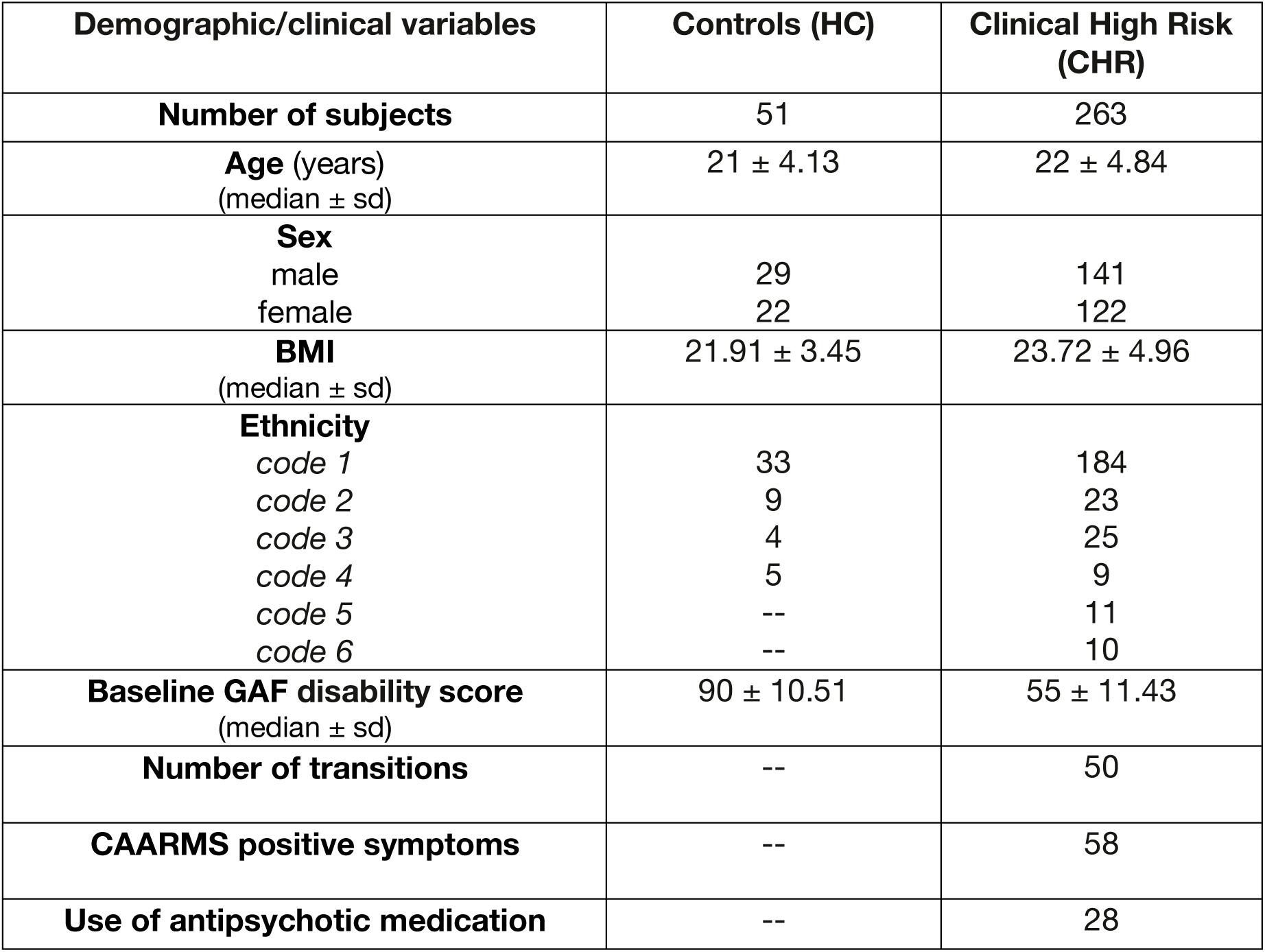
Demographic characteristics of the study population.

### Lipidomic analysis

Non-fasting serum samples, stored at −80 °C, were analyzed using an established global lipid profiling protocol, based on ultra-high pressure liquid chromatography (UHPLC) coupled to high-resolution quadrupole time-of-flight mass spectrometry (QTOF). Samples were prepared using a modified Folch procedure, as previously described^15^. In short, the samples (10 µL) were diluted with 10 µL saline (0.9%), then extracted with 120 µL of CHCl_3_:MeOH (2:1 v/v), containing a standard solution (2.5 µg/mL) comprising the following compounds: 1,2-diheptadecanoyl-sn-glycero-3-phosphoethanolamine (PE 17:0/17:0), *N*-heptadecanoyl-D-erythro-sphingosylphosphorylcholine (SM d18:1/17:0), *N*-heptadecanoyl-D-erythro-sphingosine (Cer d18:1/17:0), 1,2-diheptadecanoyl-sn-glycero-3-phosphocholine (PC 17:0/17:0), 1-heptadecanoyl-2-hydroxy-sn-glycero-3-phosphocholine (LPC 17:0) and 1-palmitoyl-d31-2-oleoyl-sn-glycero-3-phosphocholine (PC 16:0/d31/18:1) and cholest-5-en-3ß-yl heptadecanoate (CE 17:0), all purchased from Avanti Polar Lipids (Alabaster, AL, USA), and triheptadecanoin (TG 17:0/17:0/17:0) from Larodan (Solna, Sweden). The samples were then vortexed and allowed to stand on ice for 30 minutes. The samples were then centrifuged (9400 g, 3 minutes). 60 µL of the bottom layer was then collected and added to a glass vial and diluted with a further 60 µL of a mixture of CHCl_3_:MeOH (2:1 v/v). The samples were stored at −80 °C until further analysis.

In order to provide class-based calibration curves for quantitation (concentration levels 100, 500, 1000, 1500, 2000, and 2500 ng/ml, including 1250 ng/ml ISTD mixture) using the following lipids: 1,2-Dipalmitoyl-sn-glycero-3-phosphocholine (PC 16:0/16:0), 1,2-Disteayrol-sn-glycero-3-phosphocholine (PC 18:0/18:0), 1-palmitoyl-2-oleoyl-glycero-3-phosphocholine (PC 16:0/18:1), 1-octadecanoyl-sn-glycero-3-phosphocholine (LPC 18:0), 1-oleoyl-2-hydroxy-sn-glycero-3-phosphocholine (LPC 18:1), 1-oleoyl-2-hydroxy-sn-glycero-3-phosphoethanolamine (LPE 18:1), 1,2-distearoyl-sn-glycero-3-phosphoethanolamine (PE 18:0/18:0), 1-stearoyl-2-arachidonoyl-sn-glycero-3-phosphoinositol (PI 18:0/20:4), N-oleoyl-D-erythro-sphinganine (Cer d18:0/18:1) from Avanti Polar Lipids, 1-Palmitoyl-2-Hydroxy-sn-Glycero-3-Phosphatidylcholine (LPC 16:0), 1,2,3-Trioctadecanoylglycerol (TG 18:0/18:0/18:0) and 1,2,3-Trihexadecanoylglycerol (TG 16:0/16:0/16:0) from Larodan, and and 3β-hydroxy-5-cholestene 3-linoleate (CE 18:2) from Sigma Aldrich.

The samples were then analyzed using UHPLC-QTOF-MS instrument. The UHPLC system was a 1290 Infinity system (Agilent Technologies, Santa Clara, CA, USA) equipped with an autosampler maintained at 10 °C. The needle was washed with both a 10% DCM in MeOH and ACN:MeOH:IPA:H_2_O (1:1:1:1 v/v/v/v) with 1% HCOOH for a total of 7.5 seconds each. The solvents were delivered using a quaternary solvent and a column oven (set to 50 °C). The separation was performed on an ACQUITY UHPLC BEH C18 column (2.1 mm × 100 mm, particle size 1.7 µm, Waters, Milford, MA, USA). The flow rate was set at 0.4 ml/min throughout the run with an injection volume of 1 µL. The following solvents were used for the gradient elution: Solvent A was H_2_O with 1% NH_4_Ac (1M) and HCOOH (0.1%) added. Solvent B was a mixture of ACN:IPA (1:1 v/v) with 1% NH_4_Ac (1M) and HCOOH (0.1%) added. The gradient was programmed as follows: 0 to 2 min 35-80% B, 2 to 7 min 80-100 % B, 7 to 14 min 100% B. The column was equilibrated with a 7 min period of 35 % B prior to the next run. The mass spectrometer was a 6545 QTOF instrument (Agilent Technologies) equipped with a duel ion jet stream electrospray ion source. Nitrogen was generated using a nitrogen generator (PEAK Scientific, Renfrewshire, Scotland, UK).

Multiple QC samples were run throughout the run including blanks, pure standards, extracted standards, pooled serum samples and a reference serum sample (NIST 1950, National Institute of Standards, USA). The relative standard deviations (% RSD) for the peak areas of the internal standards in all standards was 16.7% (raw variation), and for the retention time RSD was on average 0.17 %. The RSD for the lipid concentrations in the pooled control samples were on average 17.8% and for the retention times on average 0.39%.

The identification was carried out in pooled serum sample, and with this information, an in-house database was created with m/z and retention time for each lipid. Identification of lipids was carried out by combining MS (and retention time), MS/MS information, and a search of the LIPID MAPS spectral database^16^, and in some cases by using authentic lipid standards. MS/MS data were acquired in both negative and positive ion modes in order to maximize identification coverage. The confirmation of a lipid’s structure requires the identification of hydrocarbon chains bound to its polar moieties, and this was possible in some cases.

### Data preprocessing

MS data were processed using the open source software MZmine 2.18^17^. The following data processing steps were applied to the raw MS data: (1) Crop filtering with a m/z range of 350-1200 m/z and a retention time (RT) range of 2 to 15 minutes; (2) Mass detection with a noise level of 1000; (3) Chromatogram builder with a min time span of 0.08 minutes, minimum height of 1200 and m/z tolerance of 0.006 m/z or 10.0 ppm; (4) Chromatogram deconvolution using the local minimum search algorithm with a 70% chromatographic threshold, 0.05 min minimum RT range, 5% minimum relative height, 1200 minimum absolute height, a minimum ration of peak top/edge of 1.2 and a peak duration range of 0.08 - 5.0 minutes; (5) Isotopic peak grouper with a m/z tolerance of 5.0 ppm, RT tolerance of 0.05 minute, maximum charge of 2 and with the most intense isotope set as the representative isotope; (6) Join aligner with m/z tolerance of 0.009 or 10.0 ppm and a weight of 2, RT tolerance of 0.1 minute and a weight of 1 and with no requirement of charge state or ID and no comparison of isotope pattern; (7) Peak list row filter with a minimum of 43 peaks in a row (10% of the samples); (8) Gap filling using the same RT and m/z range gap filler algorithm with an m/z tolerance of 0.009 m/z or 11.0 ppm; (9) Identification of lipids using a custom database (based on UHPLC-MS/MS data using the same lipidomics protocol, with RT data, MS and MS/MS) search with an m/z tolerance of 0.009 m/z or 10.0 ppm and a RT tolerance of 0.1 min.; (10) Normalization using internal standards (PE 17:0/17:0, SM d18:1/17:0, Cer d18:1/17:0, LPC 17:0) TG 17:0/17:0/17:0 and PC 16:0/d30/18:1) for identified lipids and closest internal standard (based on RT) for the unknown lipids, followed by calculation of the concentrations based on lipid-class calibration curves; (11) Imputation of missing values by half of the row’s minimum.

### Statistical methods

At the baseline, the lipidomics dataset was divided into two study groups, (1) individuals at clinical high-risk for psychosis (CHR, N=263) and (2) healthy controls (HC, N=51). Subsequently, during the follow-up period, some CHR subjects underwent symptomatic remission, such that they no longer met clinical criteria for the CHR state (CHRr), some has persistent symptoms of the CHR state (CHRp), and others transitioned to psychosis (CHRt) (**Figure 1A**).

**Figure 1.**
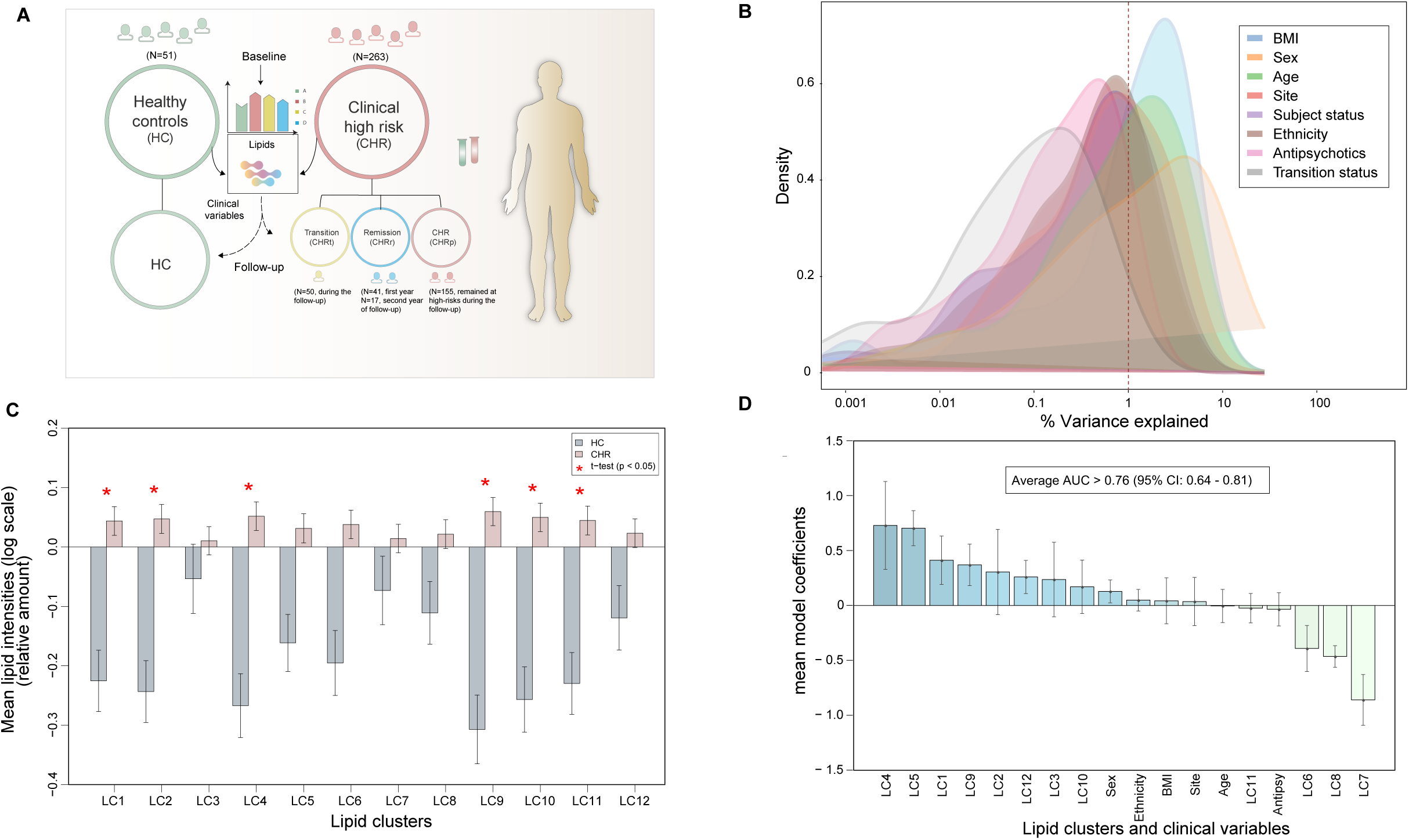
Study overview. (A) Study design and description. (B) Contributing factors affecting the variance of the lipidome. % of explained variances (EV) of all the factors across the lipid samples are shown. (C) Lipid clusters (LCs) and their mean differences between CHR and HC groups. *denotes statistically significant differences (two samples T-test, p<0.05) of group means. Lipid types within each cluster are shown in **Table 2**. (D) Model coefficients of different LCs and demographic variables for classifying CHR *vs*. HC. ‘Antipsy’ denotes antipsychotics.

The data were log2 transformed. Homogeneity of the samples were assessed by principal component analysis (PCA)^18^ and no outliers were detected. The R statistical programming language (version 3.6.0)^19^ was used for data analysis. Principal component analysis (PCA) was performed using *‘prcomp’* function included in the *‘stats’* package. *‘Heatmap*.*2’, ‘boxplot’, ‘beanplot’*, ‘*gplot*’, and ‘*ggplot2*’ libraries/packages were used for data visualization.

The effect of different factors such as age, sex, BMI and other demographic variables on the lipidomics dataset were evaluated. The data were centered to zero mean and unit variance (autoscaled). The relative contribution of each factor to the total variance in the dataset was estimated by fitting a linear regression model, where the normalized intensities of metabolites were regressed to the factor of interest, and thereby median marginal coefficients (R^2^) were estimated. This analysis was performed using the *‘Scater’* package.

Clustering of the lipidomic data was applied by using the mclust R package (version 5.4.5)^20^. Mclust is a model-based clustering method, where the model performances are evaluated by the Bayesian Information Criterion (BIC). Generally, the model with the highest BIC is chosen. At first, lipidomic clusters were generated for the HC. The mean value of the lipids in a cluster was estimated. Similarly, clustering was performed on the lipidome from the total CHR sample, and the mean value of the lipids in each cluster was estimated. A two-sample t-test was performed to assess the significance of the mean difference in the intensities of the lipids between the HC and CHR groups, within each cluster. Differences in the intensities of the lipids between the CHR subgroups and HC were assessed using analysis of variance (ANOVA), at a significance threshold of p < 0.05.

Pairwise sparse partial least squares discriminant analysis (sPLS-DA)^21^ models, comparing the lipid intensities of CHR *vs*. HC at baseline and follow-up, were developed and Variable Importance in Projection (VIP) scores^22^ were calculated. sPLS-DA modeling was performed using the ‘*splsda’* function coded in the *‘mixOmics v6*.*3*.*2’* package. Moreover, the sPLS-DA models were cross-validated^23^ by 7-fold cross-validation and models diagnostics were generated using *‘perf’* function.

The multivariate analysis was complemented by univariate analysis (two sample t-test) using the *‘t*.*test’* function, in order to identify differences in the concentration of individual lipids between CHR *vs*. HC groups, at baseline and follow-up. The consensus approach enabled us to select the altered lipid species with higher confidence. All lipids that passed the criteria of variable selection from both multi- and univariate approach, *i*.*e*., with sPLS-DA model (area under the Receiver Operating Curve (ROC), AUC >= 0.6, regression coefficients, RC (>± 0.05), VIP scores^22^ > 1) and T-test; p-value < 0.05, were listed as significant. P-values were subjected to False Discovery Rates (FDR) adjustment using ‘*p-adjust’*.

The ‘*qpgraph’* R package (version 2.18.0) was used for partial correlation network analysis. The *‘qpNrr’* function was used to estimate the non-rejection rates (NRRs) of the correlation between the lipids and/or the clinical variables. About 10,000 tests for non-rejection were performed for each pair of variables included in the correlation matrix. This analysis was performed separately for the HC, CHRp, CHRt and CHRr groups. All the spurious correlations/associations with (NRR ≤ 0.5) were removed. The Spearman’s rank correlation coefficient was estimated using ‘*rcorr’* function coded in the ‘*Hmisc v4*.*2-0’* package.

In order to understand the relative importance of lipid clusters, clinical and demographic variables in stratifying HC and CHR subjects at the baseline, we performed logistic ridge regression (LR) modelling^24^. Here, 70% of the lipidome data were used to train the model and 30% was used as a test data. LR modelling was performed using *‘cv*.*glmnet’* function deployed in R-package *‘glmnet v2*.*0-18’*. The dataset was sampled for 10,000 times. All the LR models with AUC > 0.60 (with 10-fold cross-validation) were considered. The mean AUC, log odds ratios and standard error for each variable were estimated.

LR models were then developed to stratify CHR *vs*. HC, CHRt *vs*. (CHRp + CHRr) and CHRr *vs*. (CHRp + CHRt). Twenety-one significantly changed lipids (p < 0.05) between HC, CHRp, CHRt, CHRr were used either singly or in combination for LR modelling. A recursive feature elimination scheme was implemented for the optimal selection of the lipids. The lipids in LR models were either incorporated or removed in an iterative manner, starting with all 21 lipids (p <0.05). The models were adjusted for age, gender and BMI. Accuracy of prediction was determined by AUCs. The mean AUC of a model was estimated by bootstrapping, *i*.*e*. 1000 times re-sampling with replacements and partitioning (training (70%) and test (30%) sets) the lipidomic dataset using ‘*createDataPartition’* function coded in *‘caret 6*.*0*.*84’* package. The model with the highest mean AUC was considered to be the best model which was assessed by their Receiver Operating Characteristic (ROC) Curves using *‘pROC 1*.*15*.*3’* package. Regularized ridge models in *‘cv*.*glmnet’* requires a hyper-parameter ‘λ’. Here, λ_minimum_ that corresponds to the minimum cross-validation (CV) error was determined by 10-fold CV.

To identify differences in the serum lipid levels between the CHR individuals with high or low Global Assessment of Functioning (GAF) scores^14^ at follow-up, we divided the CHR group into two subgroups with either high GAF (>65) or low GAF (≤ 65) at follow-up, corresponding to a relatively good or poor level of functioning, respectively. Welch’s t-test (p < 0.05) was used to identify the mean differences in the lipid levels. The significantly altered lipids were then subjected to iterative LR modelling.

## Results

### Lipid signature of CHR group

Lipidomic analysis, using UHPLC-MS, was performed on baseline serum samples from clinically high-risk individuals (CHR; n = 263) and healthy controls (HC; n = 51) (**Figure 1A**; **Table 1**). A total of 173 identified (based on matched UHPLC-MS/MS spectra) lipids were included in the final lipidomic dataset. Among these lipids, the following classes were represented: cholesterol esters (CEs), ceramides (Cers), lysophosphatidlycholines (LPCs), lysophosphatidylethanolamines (LPEs), phosphatidlycholines (PCs), phosphatidylethanolamines (PEs), phosphatidylinositols (PIs), sphingomyelins (SMs), and triacylglycerols (TGs).

As expected, the variables that explained most of the lipid variance were BMI, sex, and age (**Figure 1B**). Therefore, the confounding effects of these variables were taken into account in subsequent analyses. The effect of anipsychotics on the baseline lipids levels was, however, minimal, contributing to under 0.5% of explained variance (EV).

Due to the high degree of co-regulation between lipids within the same structural class, we analysed the lipidomics data using model-based clustering, which generated 12 lipid clusters (LCs) (**Table 2**). At baseline, the levels of LCs 1, 2, 4, 9, 10, and 11 were signficantly higher in the CHR group than in HC (**Figure 1C**). Clusters 2, 9, 10, and 11 comprised TGs, while LC1 contained a mixture of CEs, ceramides, PCs and PI, and LC4 included PCs and some SMs.

**Table 2.** Lipid types in each cluster at the baseline.

| Cluster Name | Cluster size | Lipid type(s) | Representative lipids in the cluster |
| --- | --- | --- | --- |
| LC1* | 24 | CE, Cer, PC, PI | CE(20:3), PC(40:5), PI(18:0/20:4) |
| LC2 | 10 | TG | TG(16:0/18:2/22:6),TG(54:7),TG(56:8) |
| LC3 | 9 | Cer, TG | Cer(d18:1/18:1), TG(50:2), TG(56:6) |
| LC4 | 18 | PCs & SM | PC(16:0/16:0), SM(d34:1), SM(d41:1) |
| LC5 | 11 | LPC, LPE | LPC(20:3), LPC(16:1), LPE(18:1) |
| LC6 | 22 | PCs, SM | PC(16:0/18:1), SM(d18:2/14:0),SM(d32:1) |
| LC7 | 22 | PC, PE | PC(O-22:2/22:3),PC(O-40:5),<br>PE(P18:0/22:6) |
| LC8 | 5 | PE | PE(16:0/18:1), PE(34:2), PE(38:6) |
| LC9 | 16 | TG | TG(14:0/16:0/18:1),<br>TG(14:0/18:1/18:1),TG(48:3) |
| LC10 | 23 | TG | TG(18:1/18:1/16:0), TG(54:2), TG(56:2) |
| LC11 | 9 | TG | TG(18:0/18:0/18:0), TG(51:2), TG(55:5) |
| LC12 | 4 | TG | TG(O-50:1) or TG(P-50:0), TG(O-52:1) or<br>TG(P-52:0), TG(O-52:2) or TG(P-52:1) |

Combining the LC data and the clinical variables, a logistic regression model was built which discriminated between CHR and HC at baseline, with AUC = 0.76 (95% CI: 0.643 - 0.816). Based on the mean model coefficients, LCs 4 and 5 had the greatest impact on the separation of CHR and HC groups, whereas demographic variables such as ethnicity, BMI, study site, antipsychotics or age had relatively little effect. Sex was the only demographic factor that had a positive impact on the regression model (**Figure 1D)**. A partial correlation analysis between various demographic variables and lipid clusters in CHR and HC subgroups is shown in **Supplementary Figure 1**.

At the individual molecular lipid level, in line with the findings at the lipid cluster level, the lipids that passed the threshold test for significance in both the multivariate and univariate tests were higher in the CHR group than in HC (**Figure 2A**), with over half being TGs. Most of the TGs that were at higher levels in the CHR group had a low carbon number and double bond count. In contrast, levels of several TGs containing longer and polyunsaturated fatty acyl chains were lower in CHR subjects than HC (**Figure 2B**).

**Figure 2.**
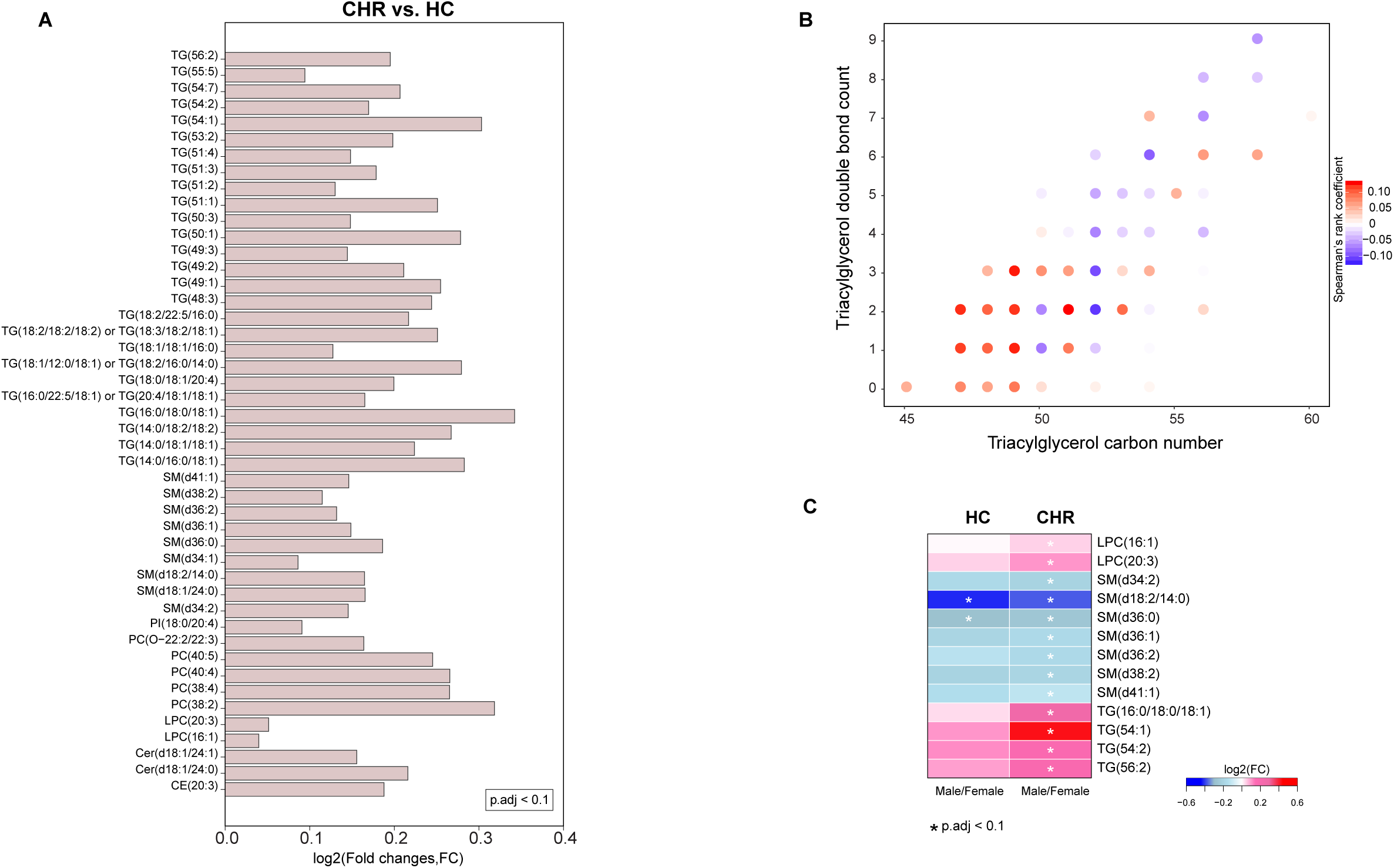
Molecular lipids in CHR. (A) Lipids selected by univariate and multivariate analysis, comparing CHR *vs*. HC. (B) Scatter plot showing the number of acyl carbons and double bonds in individual TGs. The spearman coefficient (δ) is color coded with red as positive, and blue as negative correlation between the TG levels in CHR vs. HC. (C) List of lipids that are significantly altered between CHR vs. HC as well as between males and females within each group at baseline. The abbreviation for lipids cholesterol esters (CEs), ceramides (Cers), lysophosphatidlycholines (LPCs), lysophosphatidylethanolamines (LPEs), phosphatidlycholines (PCs), phosphatidylethanolamines (PEs), phosphatidylinositols (PIs), sphingomyelins (SMs), and triacylglycerols (TGs).

As there was a strong effect of sex in the logistic regression model, we also examined differences in relation to sex. There were 13 lipids which differed by both sex and by group (CHR *vs*. HC) (**Figure 2C** and **Supplementary Figure 2A-I**). The majority of these were SMs, all of which were at lower levels in male CHR individuals (**Figure 2C** and **Supplementary Figure 2D-F**). Conversely, levels of four TGs, were higher in male CHR individuals (**Figure 2C** and **Supplementary Figure 2G-I**).

### Lipid signatures of clinical outcomes in CHR subjects

The same 12 lipid clusters described for the CHR vs HC comparisons (above) were used to compare the CHR outcome subgroups and HC. Five lipid clusters significantly differed between these subgroups at baseline. For 4 of these clusters (LC1, LC2, LC9 and LC10), the subgroups that contained subjects who had persistent symptoms or were psychotic at follow-up (CHRp and CHRt) had higher levels than the subgroups that contained subjects who were in remission (CHRr) or were HC; **Table 2**; **Figure 3A**).

**Figure 3.**
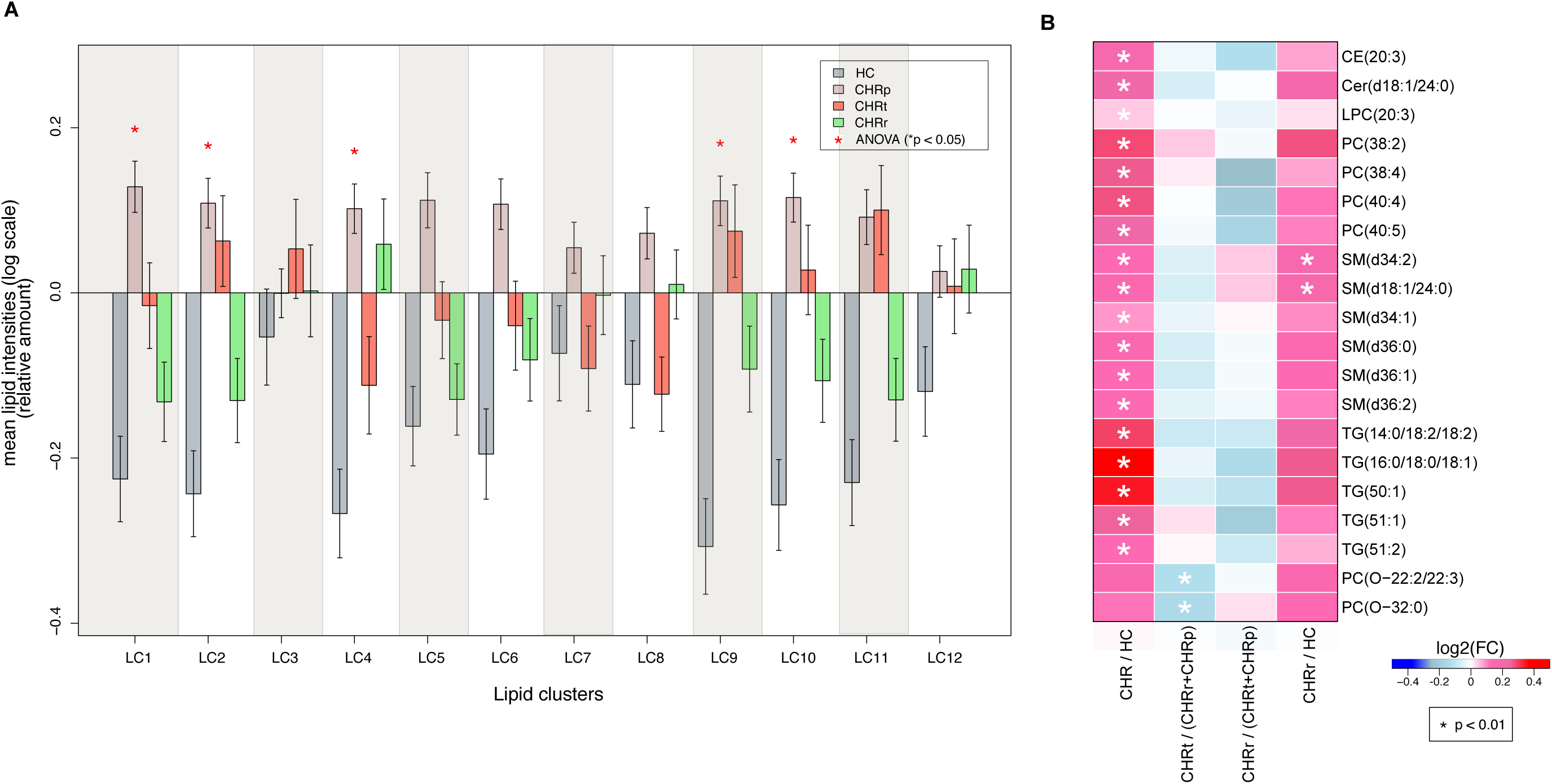
CHR and health outcomes in the follow-up. (A) Lipid clusters at baseline for four study groups: CHRp (remained CHR in the follow-up), transition from CHR to psychosis (CHRt), remission from CHR (CHRr) and HC. *denotes statistically significant differences (ANOVA) of the group means. (B) List of lipids that were different between these groups. FC stands for fold change.

In terms of individual lipids, there was a trend for lower levels in the CHRt subgroup than in the CHR subgroups containing subjects who did not become psychotic (CHRs + CHRr) (**Figure 3B**). Among these lipids, the levels of two ether phospholipids, PC(O–22:2/22:3) and PC(O–32:0) were significantly lower in the CHRt than in the CHRr and CHRp subgroups (**Figure 3B**). Two lipids, SM(d34:2) and SM(d18:1/24:0), had higher levels in the CHRr subgroup than in the HC group.

### Lipids as a predictor in CHR for psychosis

Next, we examined whether the lipids that significantly differed (p < 0.05) between the CHRp, CHRt and CHRr subgroups could be used to stratify individuals within the total CHR sample. These lipids were subjected to LR modelling in an iterative manner and optimal sets of lipids were identified. Cer(d18:1/24:0), LPC(22:5), PC(38:4), PC(40:5), PC(O-32:0) were able to differentiate CHRt from CHR subjects who did not become psychotic (mean AUC = 0.807, 95% CI = 0.685-0.928). Ether phospholipid PC(O-32:0) was diminished in CHRt in comparison to CHR individuals that did not develop psychosis (**Figure 4E**). CHRr were distinguished from CHR subjects who did not achieve remission by Cer(d18:1/24:0), LPC(22:5), PC(38:4), PC(40:5), PC(O-32:0) and SM(d18:1/24:0) (mean AUC = 0.833, 95% CI = 0.718-0.947) (**Figure 4B-C**). Cer(d18:1/24:0), LPC(22:5), PC(38:4), PC(40:5), PC(O-32:0), SM(d18:1/24:0), SM(d36:0) and SM(d36:1) we were able to distinguish the total sample of CHR individuals from HC with a good level of accuracy (mean AUC = 0.828, 95% CI = 0.705-0.951) (**Figure 4A**).

**Figure 4.**
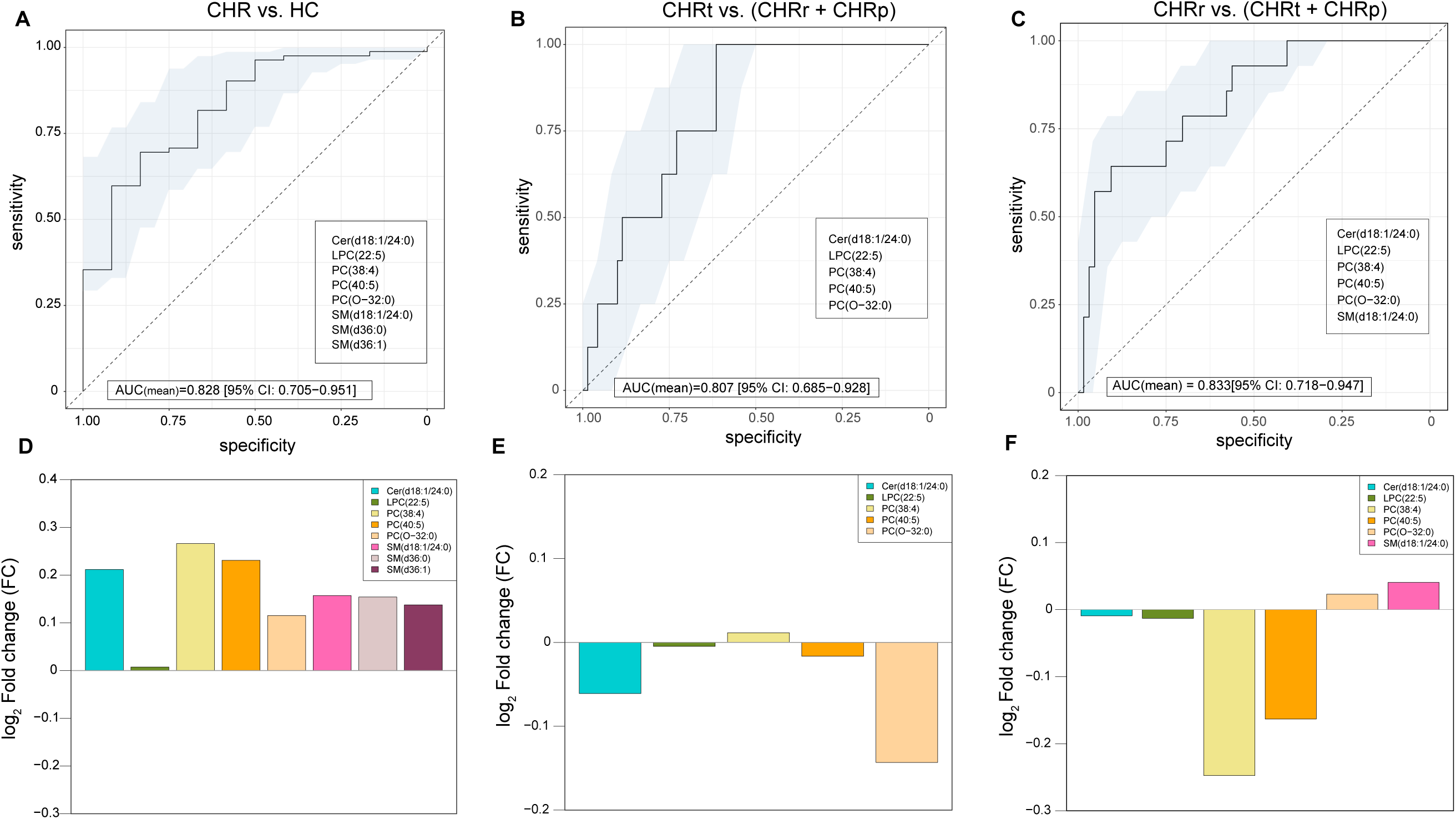
Predictive models of CHR conditions. Logistic ridge regression (LR) models showing lipids as predictive markers to stratify patient groups of HCs and/or CHR (divided into CHRp, CHRt and CHRr). (A-C) Receiver-operating characteristic (ROC) curves showing the performance of the LR models (CHR *vs*. HC, CHRt *vs*. (CHRp + CHRr) and CHRr *vs*. (CHRp + CHRt)) with highest mean AUCs. The light blue shaded area denotes the 95% confidence intervals (CI) as calculated using bootstrapping. Specific lipids that comprise each of these models are shown. (D-F) Log2 fold change (FC) in the intensities of the lipids corresponding to the LR models.

### Association between lipids and functional outcomes

There were significant differences (Welch’s t-test, p < 0.05) in baseline serum lipids between CHR subjects with high and low GAF disability scores at follow up: levels of triacylglycerols TG(52:2), TG(52:3), TG(52:4) and TG(56:6) were markedly higher (p < 0.05) in the subjects with low GAF disability scores (**Supplementary Figure 3**). Iterative LR modelling of these significantly altered TGs identified (TG(52:3), TG(52:4) and TG(56:6)) as an optimal set for distinguishing between subjects with high and low GAF disability scores at follow-up (mean AUC = 0.797, 95% CI = 0.68-0.90; **Supplementary Figure 4**).

## Discussion

Our first major finding was that the blood levels of many lipids were higher in people at CHR for psychosis than HC. The types of lipid that were different were similar to those previously seen in comparisons of patients with psychosis and HC^25,26^. The identification of lipidomic differences before the onset of psychosis is consistent with similar findings in healthy children^27^ who subsequently reported psychotic experiences in later life, in siblings of psychotic patients^28^, and suggests that altered serum lipid levels are related to an increased vulnerability to psychosis, and are not simply a secondary consequence of the disorder or its treatment.

The higher levels of TGs in the CHR group were primarily in TGs with low acyl carbon number and double bond count (*i*.*e*., those containing shorter and more saturated fatty acyl chains). A similar TG lipid signature was previously observed in a subgroup of FEP patients who rapidly gained weight after presentation^11^. Serum TGs with low acyl carbon number and double bond count are also increased in NAFLD^10^ and precede clinical type 2 diabetes^29^. In NAFLD, these TGs characterize the subtype of NAFLD associated with obesity and insulin resistance^30^. At least in part, changes in these TGs may be related to increased *de novo* hepatic lipogenesis, a hallmark of NAFLD^31, 32^.

When comparing the three CHR sub-groups at the lipid cluster level (**Figure 3**), individuals whose symptoms remitted (CHRr) had a similar lipid profile to HC. Although the lipids tended to be higher in CHRr than in HC, the only significant difference at the individual lipid levels was in two SMs, SM(d34:2) and SM(d18:1/24:0), with these two lipids being elevated also in whole CHR as compared to HC. These SMs were previously found to be associated with obesity and insulin resistance^33^, and SM is considered a risk factor of coronary artery disease^34^. Altered SM levels as compared to HC in CHRr may thus reflect impared metabolic profile in CHR individuals.

Using a machine learning approach, we developed diagnostic signatures that discriminated subgroups within the CHR sample with distinct clinical outcomes. The relatively high accuracy of the model suggests that measures of blood lipids may help clinicians predict outcomes in this population. CHR individuals who later developed psychosis were discriminated from those who did not by lower levels of ether phospholipids. Ether phospholipids are highly enriched in brain^35^ and have many structural and functional roles^36^. Plasmalogens are a structural subgroup of ether phospholipids that are supplied to the brain by the liver^37^. They thought to be scavengers of free radicals, and may act as endogenous antioxidants^38, 39^. Our data thus raise the possibility that CHR individuals who develop psychosis may have an increased vulnerability to oxidative stress. This is of particular interest, as an independent body of work has implicated oxidative stress in the pathophysiology of psychosis^40^. We have also collected measures of oxidative stress from the present sample and will assess the relationship between these and ether phospholipids in a forthcoming study.

An unexpected finding was that within the total CHR sample, there were marked sex differences in lipid profile. Although the trend of lipid changes, when comparing CHR and HC individuals, was the same for both sexes, sex differences were much more pronounced in the CHR group than in HCs. While TGs and LPCs were increased in males in CHR, SMs were decreased. Plasma SMs are known to be elevated in females^41^, which reflects the effect of 17 beta estradiol on serum SM levels^42^. This finding suggests that sex may be an important confounder that needs to be considered in studies of lipid metabolism in psychosis.

In conclusion, although the mechanisms linking dysregulation of lipid metabolism with the pathophysiology of psychosis is unclear, our findings suggest that metabolic abnormalities are evident in people who are vulnerable to psychosis, but do not have the disorder and have not been treated. A similar lipid profile is observed in patients with NAFLD and in pre-diabetes, as well as in non-obese FEP patients who later gain weight. Secondly, our data also suggest that assessment of the circulating lipidome may assist in the identification of CHR individuals at highest risk of transition to psychosis. An AUC of 0.83 for predicting transition is promising, but the model must be validated in similar independent CHR datasets. The predictive power of lipidomic data may be enhanced by combining these with measures of other factors that may influence clinical outcomes in CHR subjects, such as neuroimaging data, psychopathology, oxidative stress, proteomic, and inflammatory markers^43^.

## Data Availability

Reasonable requests for access to the data of this study will be carefully considered by the EU-GEI High Risk Study publication committee.

## Acknowledgements

The authors thank to Cecilia Carlsson for technical assistance in lipidomic analysis.

## Author contributions

P.M. and M.O. initiated, designed, and supervised the study. N.B.-V., C.I., M.N., T.P., A.R.-R., S.R., G.S., R.B., M.-O.K., L.d.H., M.v.g.G., L.V., and P.M. conducted patient recruitment and follow-up in the EU-GEI study. T.H. acquired lipidomics data by mass spectrometry. A.D., P.S., M.J.K., and M.O. analyzed the data. A.D., P.S., and M.O. wrote the first draft of the manuscript. All authors edited the manuscript and approved the final version.

## EU-GEI High Risk Study Group Author

Philip McGuire^1^

Lucia R. Valmaggia^2^

Matthew J. Kempton^1^

Maria Calem^1^

Stefania Tognin^1^

Gemma Modinos^1^

Lieuwe de Haan^3,4^

Mark van der Gaag^5,6^

Eva Velthorst^3,7^

Tamar C. Kraan^3^

Daniella S. van Dam^3^

Nadine Burger^6^

Barnaby Nelson^8^

Patrick McGorry^8^

G Paul Amminger^8^

Christos Pantelis^8^

Athena Politis^8^

Joanne Goodall^8^

Anita Riecher-Rössler^9^

Stefan Borgwardt^9^

Charlotte Rapp^9^

Sarah Ittig^9^

Erich Studerus^9^

Renata Smieskova^9^

Rodrigo Bressan^10^

Ary Gadelha^10^

Elisa Brietzke^11^

Graccielle Asevedo^10^

Elson Asevedo^10^

Andre Zugman^10^

Neus Barrantes-Vidal^12^

Tecelli Domínguez-Martínez^13^

Anna Racciopi^14^

Thomas R. Kwapil^15^

Manel Monsonet^14^

Araceli Rosa^16^

Ariel Frajerman^17^

Boris Chaumette^17^

Julie Bourgin^17^

Oussama Kebir^17^

Célia Jantac^17^

Marie-Odile Krebs^17^

Dorte Nordholm^18^

Lasse Randers^18^

Kristine Krakauer^18^

Louise Glenthøj^18^

Birte Glenthøj^19^

Merete Nordentoft^18^

Stephan Ruhrmann^20^

Dominika Gebhard^20^

Julia Arnhold^21^

Joachim Klosterkötter^21^

Gabriele Sachs^22^

Iris Lasser^22^

Bernadette

Winklbaur^22^

Philippe A Delespaul^23,24^

Bart P. Rutten^23^

Jim van Os^1,23^

## AFFILIATIONS OF GROUP AUTHORS

1. Department of Psychosis Studies, Institute of Psychiatry, Psychology & Neuroscience, King’s College London, De Crespigny Park, Denmark 458 Hill, London, United Kingdom SE5 8AF.
2. Department of Psychology, Institute of Psychiatry, Psychology & Neuroscience, King’s College London, De Crespigny Park, Denmark Hill, 456 London, United Kingdom SE5 8AF.
3. AMC, Academic Psychiatric Centre, Department Early Psychosis, Meibergdreef 5, 1105 AZ Amsterdam, The Netherlands.
4. Arkin Amsterdam, The Netherlands.
5. VU University, Faculty of Behavioural and Movement Sciences, Department of Clinical Psychology and EMGO+ Institute for Health and Care Research, van der Boechorststraat 1, 1081 BT Amsterdam, The Netherlands.
6. Parnassia Psychiatric Institute, Department of Psychosis Research, Zoutkeetsingel 40, 2512 HN The Hague, The Netherlands.
7. Icahn School of Medicine at Mount Sinai, department of Psychiatry, 1425 Madison Ave, New York, NY 10029.
8. Centre for Youth Mental Health, University of Melbourne, 35 Poplar Road (Locked Bag 10), Parkville, Victoria 485 3052, Australia.
9. University Psychiatric Hospital, Wilhelm Klein-Strasse 27, CH-4002 Basel, Switzerland.
10. LiNC - Lab Interdisciplinar Neurociências Clínicas, Depto Psiquiatria, Escola Paulista de Medicina, Universidade Federal de São Paulo – UNIFESP.
11. Depto Psiquiatria, Escola Paulista de Medicina, Universidade Federal de São Paulo – UNIFESP.
12. Departament de Psicologia Clínica i de la Salut (Universitat Autònoma de Barcelona), Fundació Sanitària Sant Pere Claver (Spain), Spanish Mental Health Research Network (CIBERSAM).
13. CONACYT-Dirección de Investigaciones Epidemiológicas y Psicosociales, Instituto Nacional de Psiquiatría Ramón de la Fuente Muñiz (México).
14. Departament de Psicologia Clínica i de la Salut (Universitat Autònoma de Barcelona).
15. Department of Psychology, University of Illinois at Urbana-Champaign (USA).
16. Departament de Biologia Evolutiva, Ecologia i Ciències Ambientals (Universitat de Barcelona), Spanish Mental Health Research Network (CIBERSAM).
17. University Paris, GHU Paris Sainte-Anne, C’JAAD, Inserm U1266, Institut de Psychiatrie (CNRS GDR 3557) Paris, France
18. Mental Health Center Copenhagen and Center for Clinical Intervention and Neuropsychiatric Schizophrenia Research, CINS, Mental Health Center Glostrup, Mental Health Services in the Capital Region of Copenhagen, University of Copenhagen.
19. Centre for Neuropsychiatric Schizophrenia Research (CNSR) & Centre for Clinical Intervention and Neuropsychiatric Schizophrenia Research (CINS), Mental Health Centre Glostrup, University of Copenhagen, Glostrup, Denmark
20. Department of Psychiatry and Psychotherapy, Medical Faculty and University Hospital, University of Cologne, Cologne, Germany.
21. Psyberlin, Berlin, Germany.
22. Medical University of Vienna, Department of Psychiatry and Psychotherapy.
23. Department of Psychiatry and Neuropsychology, School for Mental Health and Neuroscience, Maastricht University Medical Centre, P.O. Box 616, 6200 MD 464 Maastricht, The Netherlands
24. Mondriaan Mental Health Trust, P.O. Box 4436 CX Heerlen, The Netherlands

